# Analysis of the Worldwide Corona Virus (COVID-19) Pandemic Trend; A Modelling Study to Predict Its Spread

**DOI:** 10.1101/2020.03.30.20048215

**Authors:** Muhammad Qasim, Waqas Ahmad, Minami Yoshida, Maree Gould, Muhammad Yasir

## Abstract

**Objective:** The Coronavirus (COVID-19) has advanced into 197 countries and territories leaving behind a total of 372,757 confirmed cases and 16231 deaths.

**Methods:** One the basis of WHO situation reports data of COVID-19 along with daily official reports from the Japan, China and the Kore we modelled the spread of COVID19 by using the Successive Approximation Method. We defined the two state of data to find the mean ratio (η) of the present cases count to the sum of previous and present cases. This ratio further predicts the future state of COVID-19 pandemic.

**Results:** The mean ratio (η) of expected cases were found 0.485, while the mean ratio for deaths was found to be 0.49. We calculated worldwide expected lower bound value for confirmed cases 247007 cases with maximum limit of 1667719 cases and minimum deaths count 8660 with upper limit of 117397 deaths in next 30 days. While in the case of Iran, a large increase in the number of deaths are expected in the upcoming 30 days with lower bound value of 1140 deaths and maximum value of 598478 deaths.

**Interpretation:** Iran whole population is on risk.

## Introduction

On the 31^st^ of December 2019, WHO was alerted to a group of pneumonia cases of unknown origin in Wuhan City, Hubei Province, China. Through throat swab samples the causative agent was identified by the Chinese Centre for Disease Control and Prevention (CCDC) on the 7 ^th^ of January 2020. While on the 12^th^ of January, the CCDC shared genome sequence of a causative agent of corona virus disease 2019. The virus was later named Severe Acute Respiratory Syndrome Coronavirus 2 (SARS-CoV-2) and the disease was renamed COVID-19 by the World Health Organization (WHO)^1^

According to WHO, the virus has spread to 197 countries and territories and infected 372,757 people and caused 16,231 deaths (4.35 % mortality rate).^2^ This mortality rate of COVID-19 varies with the addition of new cases.^3^ As compared to other two types of Corona virus Severe Acute Respiratory syndrome (SARS)^4^ and Middle East Respiratory Syndrome (MERS)^5^ which has mortality rate of 9.6% and 34.45 respectively the COVID-19 current mortality rate is almost 4.14%. The average incubation periods for COVID-19 has been reported 4-5 days with an average time of 14 days^6 7 8^ from start symptoms to death which is almost similar to MERS.^9^ Since human to human transmission of COVID-19 has been confirmed, respiratory droplets and contact are treated as the main path of its transmission so its patients need to quarantine to reduce its spread.^10^ A recent study confirmed the spread of COVID-19 through aerosol droplets as SARS-CoV-2 remained viable in aerosols for 3 hours with little reduction in infectious titre from 10^3.5^ to 10^2.7^ 50% tissue-culture infectious dose (TCID_50_) per litre of air and its reduction rate has been found similar to SARS-CoV-1, from 10^4.3^ to 10^3.5^ TCID_50_ per millilitre. While COVID-19 was found to be more stable on plastic and stainless steel than on copper and cardboard, the viable virus was detected up to 72 hours after application to these surfaces^11^ which partly explains it lethality. Another unique characteristic of COVID-19 was that its reoccurrence was reported in an oropharyngeal swab test.^12^ Further evolution of COVID-19 has been documented and a second strain of the virus was identified and mutations seen in 149 sites across the strains and the strains have been reclassified as L and S.^13^ But this claimed was countered by Maclean et al., 2020 who diffused this information by considering it as miss interpretation of the SARS-CoV-2 data, and highlighted its methodological limitations.^14^

Since the outbreak started, the reported number of cases and deaths is increasing exponentially despite precautionary measures worldwide. Real-time data reports and forecasting are essential tools for tackling measures and policy formulations to contain the infection.^15,16^ Epidemiological predictions based on real time data create an opportunity to forecast the geographical spread of diseases and also give estimate for burden of case counts to early want to the public health system and paved path for interventions during a pandemic crisis. Exploiting all available real time data is an important aspect of modern pandemic response and helps to take strategic decisions with evidence based support.^17^

A few studies have been reported since the start of COVID-19 to model disease prevalence, spread and forecast its expansion. A modelling study conducted based upon the number of cases exported from Whuan to international locations predicted an epidemic doubling time of 6.4 days.^6^ Another model reported by the Imperial College of London estimate the potential total number of corona virus cases in Whuan city by using the total volume of international travel from Wuhan over the last two months.^18^ Another model was reported to estimate the total number of cases over Diamond Princess Cruise ship by utilizing the number of positive cases on-board at the cruise ship in Japan.^19^ To model outbreak size in Wuhan City by considering the governmental actions, individual behavioural response, zoonotic transmission and the emigration of a large proportion of the population over a short time period was developed and compared with influenza pandemic of UK in 1918.^20^ Similarly the risk of importation cases of COVID-19 to Africa from China was estimated based on flight data that originated from China and classified all of Africa into three clusters based on calculated risk.^21^ But most of these studies, have used air traffic as a base line for modelling or they consider data from only one city to predict the size of the outbreak and the ability of the country to contain the outbreak. All these different mathematical and statistical models proposed for COVID-19 have different strengths and weaknesses. In particular, some of these model specifications are better than others in distinct scenarios and in distinguishing prediction targets such as different measures of timing or the severity of the COVID-19. Additionally, there is always room for enhancing the efficiency of quantitative analyses and their results to make communication of data interpretation easy to public health concerned authorities. An accurate predictive model can help in resource mobilization and current and future disease management.

In this study, we have utilized the Successive Approximation Method by using real-time COVID-19 global data to model daily reported cases and predict the future state of the COVID- 19 pandemic. Our findings suggest that despite self-isolation, the banning of social occasions and the implementation of quarantine worldwide in major cities such as Korea, Japan and China, the mean ratio (η) of expected cases and death is still progressing. Our forecast of epidemic rolling will help realise the public health risk worldwide accounting for social, economic and health implications. For instance, the accurate and reliable predictions of COVID-19 pandemic size can help with planning how many COVID-19 vaccination doses to produce and by which date they will be required.

## Methods

Data collected from the daily situation reports of WHO^2^ from the 21^st^ of January to the18th of March 2020, daily reports from the Ministry of Health, Labour and Welfare of Japan^22^, National Health Commission of the People of China^23^ and the Korean Centre for Disease Control^24^.By using this worldwide data set, we can model the data using the Successive Approximation Method. We define the two set of data to find the mean ratio (η) of present cases count to the sum of previous and present cases. This ratio was further used to predict the future state of the expected cases and deaths counts.

We considered *y*_*n*_ is the count of confirmed reported cases at day *n*. We define the ratio of confirmed cases for two consecutive days *n* and *n −1* as:

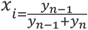

Similarly for x_i+1_ we can write the above equation as

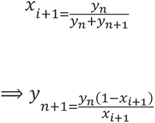

We have calculated all the x_i_ for the available data and calculated the following

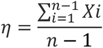

For the prediction of next sate we used x_i+1_= η to predict the expected growth of sample space (i,e. expected patients and expected deaths). The lower bound of our proposed model is calculated by selecting the maximum x_i_ between any two successive samples. It can be noted that the proposed model works when 0< x_i_ ≤1 When x_i_ → 0, the η decreases and the sample space grows exponentially.

In next step, the approximation produced can be further extended for the next day by subtracting the deaths (d) and approximating x_i_= η as

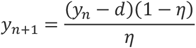

Where η_x_→ 0, the entire population is expected to become infected similarly when η_x_→ 1, the, the incidence of new patients counts significantly reduce.

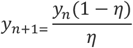

The lower bound of proposed model is calculated by selecting η=max (x_i_)

By using this mathematical model we also forecast the probable spread of COVID-19 in countries that were initially affected and calculate the expected size of cases in their cities which reported with high incidence of COVID-19 cases and deaths (supplementary file 1). The predicted number of cases and deaths reflects the potential impact of the various social and personal non-pharmaceutical interventions that have been progressively and quickly implemented in January 2020. We calculated the lower bound limit for each respective country and their capital cities. In order to evaluate the various countries strategies to contain COVID- 19 to present a model for other counties to formulate their policies to address the pandemic, we plotted the trend of pandemic for China, Japan, South Korea and Iran. We consider these countries as representative of countries with high, medium and low medical health security systems according to their ranking by the Global Health Security (GHS) Index ^25^ such as South Korea (GHS 9/195), Japan (GHS 21/195), China (GHS 51/195) and Iran (GHS 97/195 respectively.

## Results

By plotting the data according to our mathematical modelling, the mean ratio (η) worldwide has been found in 0.485 in the current state on March the19th, 2020 and the ratio gradually increased from 0.35, to remain stable for a short time in early March at 0.49. (**Figure 1a**) By using this mean ratio (η), we have plotted the expected lower bound cases and maximum upper limit of our proposed model for the next 30 days to predict the worldwide size of the pandemic in the coming days. It has been found that the lower bound count for confirmed cases for next 30 days will be 247007 and the maximum upper limit of cases will be 1667719. (**Figure 1b**). Similarly, we have plotted the expected death counts by using same mean ratio (η) and found the worldwide death count due to the pandemic with a lower bound value of 8,660 deaths and a maximum upper limit count of 117397 (0.1 million) deaths in next 30 days. (**Figure 1c**)

**Figure 1.**
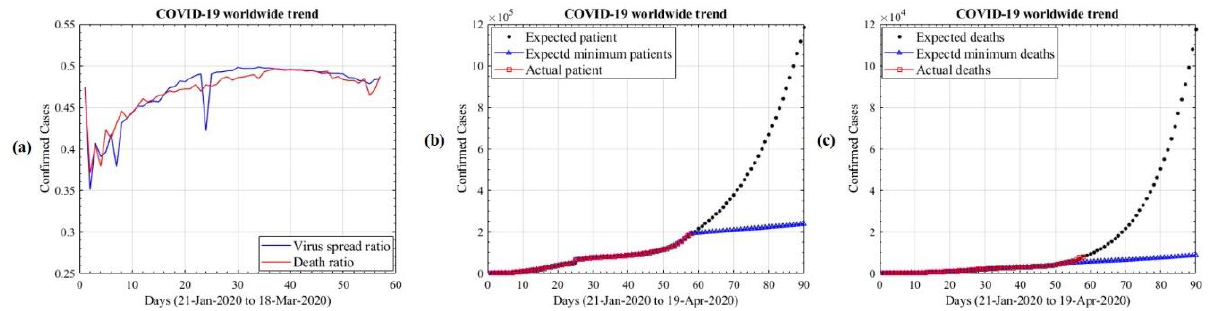
(a) Mean ratio (η) of worldwide reported case and deaths of COVID-19, (b) Prediction curves for worldwide confirmed cases of COVID19, (c) Prediction curves for worldwide deaths due to COVID19

In order to evaluate and compare the countries strategies to contain COVID-19 based on their GHS index we plotted the pandemic trends for China, Japan, South Korea and Iran in order to assist other counties and international agencies to formulate their policies to address this pandemic. In the case of South Korea, the mean ratio (η) has increased significantly from the 1^st^ case reported in the country to reach 0.48 and the country now struggles to contain COVID- 19 and to stabilise the situation in that country. Their daily case mean ratio (η) is now closing on 0.5 which indicates that the number of new cases are not increasing with respect to the previous day but their mean death rate ratio is still progressing (**Figure 2a-i**). We calculated the expected size of the pandemic for the next 30 days and the lower bound count for new cases of 11,279 and the maximum count would be 482,874 cases (**Figure 2a-ii**). As far as the current mortality rate for the Republic of Korea (ROK) was concerned which is 1.32, then the expected number of deaths will be approximately148 with respect to lower bound cases. Almost the same mean ratio (η) was found for Daegu, the main city of ROK, and now a new case mean ratio (η) almost reaching 0.5 which means that new cases are stabilised with respect to the previous day (**Figure 2a-iii**). While the predicted number of cases for Daegu city determined 7,657 with a lower bound count and a maximum value found 305,884 (0.3 million) new confirmed cases. (**Figure 2a-iv**)

**Figure 2.**
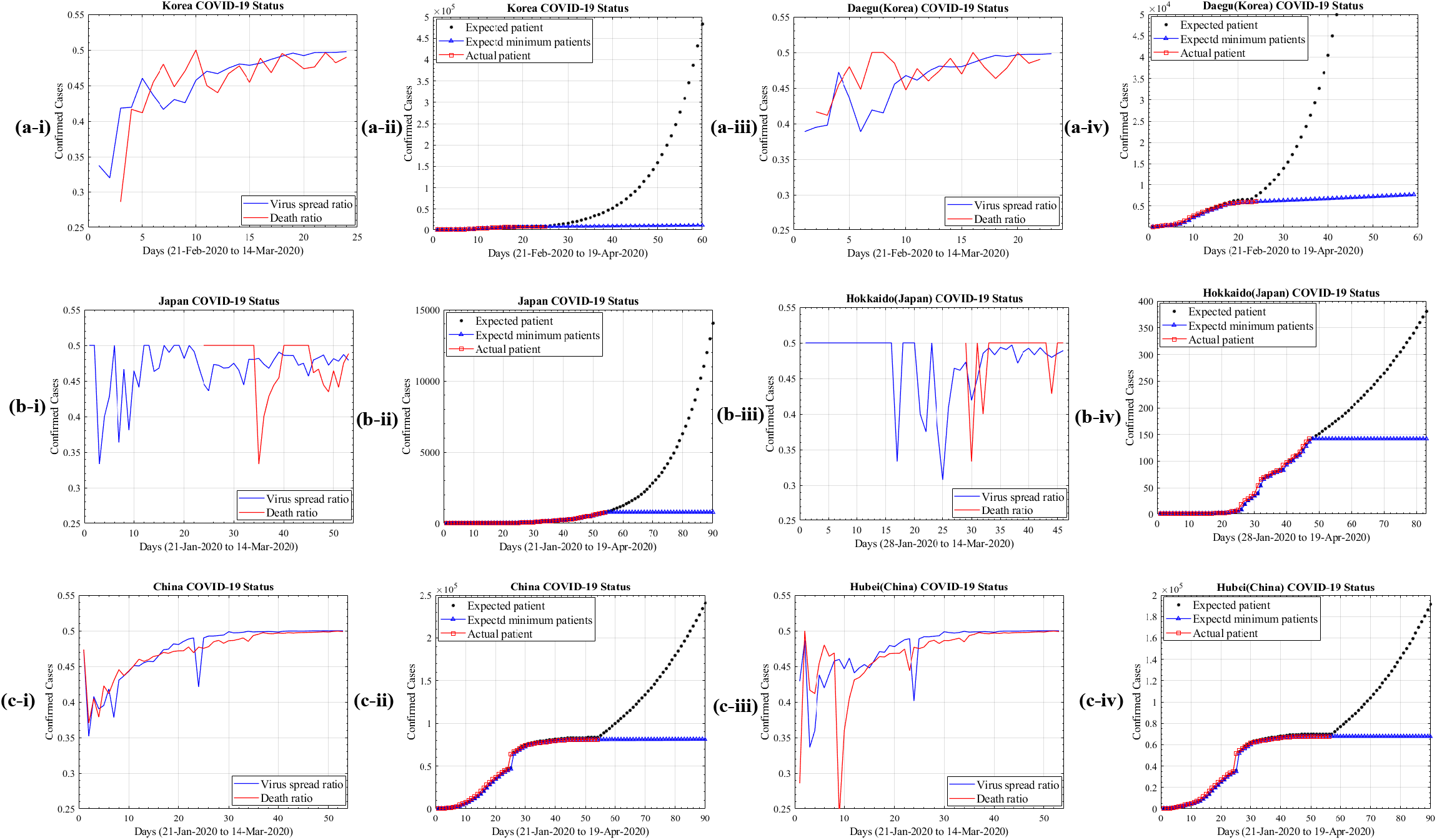
(a-i) Mean ratio (η) of Republic Of Korea reported case and deaths of COVID-19, (a- ii) Prediction curves for Republic Of Korea confirmed cases of COVID19, (a-iii) Mean ratio (η) of Republic Of Korea, Daegu City reported case and deaths of COVID-19, (a-iv) Prediction curves for Republic Of Korea, Daegu City confirmed cases of COVID19, (b-i) Figure 8 Mean ratio (η) of Japan reported case and deaths of COVID-19, (b-ii) Prediction curves for Japan confirmed cases of COVID19, (b-iii) Mean ratio (η) of Japan’s city Hakido reported case and deaths of COVID-19, (b-iv) Prediction curves for Japan’s city Hakido confirmed cases of COVID19, (c-i) Mean ratio (η) of China reported case and deaths of COVID-19, (c-ii) Prediction curves for China confirmed cases of COVID19, (c-iii) Mean ratio (η) of China’s city Hubei reported case and deaths of COVID-19, (c-iv) Prediction curves for Japan’s city Hubei confirmed cases of COVID19

In Japan, the mean ratio (η) value showed successful containment of the COVID-19 spread and death count over a long period of time with minor fluctuations. The condition stabilised (η=0.5) from time to time, both for new confirmed cases and death counts (**Figure 2b-i**). But a developing trend is showing that the number of new cases is increasing and we predicted the pandemic count for Japan in next 30 days of new cases lower bound value 781 with a maximum count value of 14,062 (**Figure 2b-ii**). The mean ratio (η) for its most affected city of Hakido provides a stabilised death count and the ratio for new cases also stabilised with a mean ratio (η) 0.5. (**Figure 2b-iii**). While expected cases count for the next 30 days of Hakido city is predicted with a lower count value of 394 and a maximum upper limit of 451 cases (**Figure 2b-iv**).

For China, the mean ratio (η) increased significantly from the 1^st^ day of new cases reported and in its current state has almost stabilised (**Figure 2c-i**). The predicted count for new cases in next 30 days found 8,1381 lower bound value and 240,951 (0.2 million) at the maximum limit. (**Figure 2c-ii**). The mean ratio (η) value trend for the city of origin of COVID-19, Hubei China showed two high outbreaks one at the end of January and another in mid-February 2020, while current trend has stabilised **(Figure 2c-iii**). While the predicted number of cases according to its mean ratio (η) were found of 67,829 with lower bound limit and a maximum upper limit of 191,787 cases would be expected within the next 30 days. (**Figure 2c-iv**).

Iran was the 2^nd^ most affected country outside of China after the outbreak and it has been found that mean ratio (η) for Iran is still increasing at 0.47 for new cases with a death mean ratio (η) of 0.45 indicating that the number of deaths will increase daily with respect to previous days, as more new cases are diagnosed with respect to the last day (**Figure 3a**). According to its mean ratio (η) the expected number of cases in next 30 days with a lower bound limit of 38538 while the maximum limit, would provide the worse possible situation as their entire population could be infected. (**Figure 3b**) The predicted value for deaths counts for Iran with lower bound count found 1140 with an upper limit of 598478 (**Figure 3c**) If the current mortality rate of Iran (7.86 %) is concerned the expected number of death would be according to predicted lower case number will be 2697 in next 30 days.

**Figure 3.**
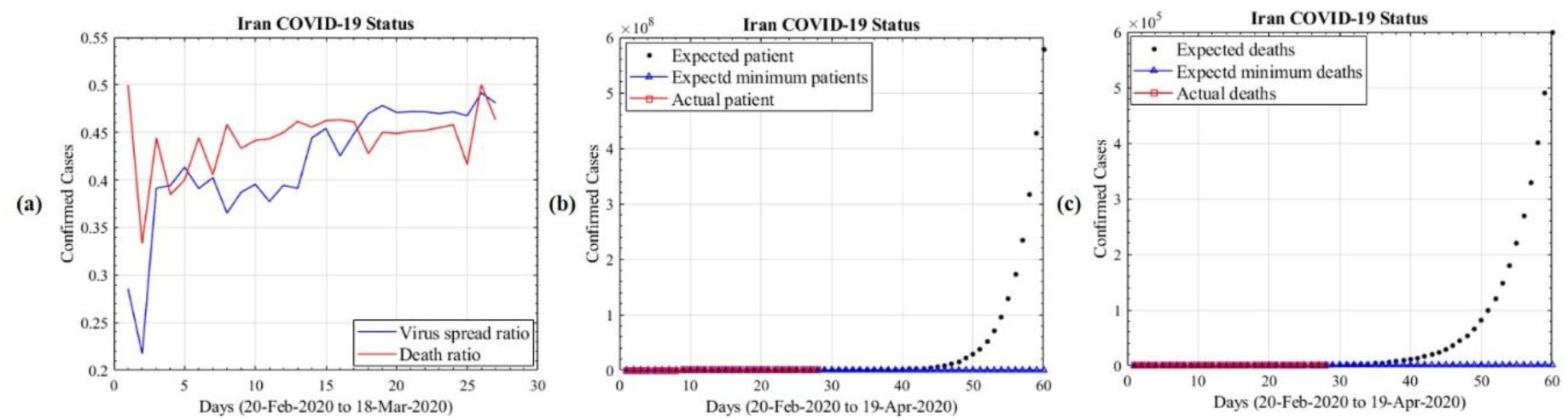
(a) Mean ratio (η) of Iran reported case and deaths of COVID-19, (b) Prediction curves for Iran confirmed cases of COVID19, (c) Prediction curves for Iran deaths due to COVID19

## Discussion

Early, rapid and large scale testing of every suspected case of COVID-19 is critical to contain and prevent its further transmission as has been emphasized by WHO.^26^ COVID-19 has been declared a pandemic and has affected 197 countries and territories to date, with a total of 372,757 confirmed cases and 16,231 deaths reported^2^ and this is increasing by the day. To predict the further spread of COVID-19 worldwide, we used mathematical modelling to draw relevant information to help international organisations and countries to plan their policies and implement measures accordingly. According to our model the mean ratio (η) for new cases in China, Republic of Korea has stabilised and they have contained the virus but still for Korea, the mean ratio (η) of the potential number of new death cases still remains at 0.48 there are more deaths expected. But the expected number of cases will be low in coming days. This shows that quarantine, social distancing, and the isolation of infected populations can successfully contain the pandemic. Despites all such measures China and Korea will still at risk as there are the possibility of reoccurrence^12^ due to the COVID-19 ability to cause reinfection in recovered patients. Similarly some cases can be still hidden and act as carrier and silent spreader. As the symptoms can present as pneumonia and have an incubation period of 14 days so it is expected that many hidden cases are waiting to resurface. But at the same time it has been reported that COVID-19 infectivity gradually decreased in the tertiary patients which is a good sign to contain further spread.^8^ Therefore, all precautionary measures are required to remain active for longer periods of time until a vaccine finally becomes available.

Our model showed that Japan successfully managed to contain the COVID-19 and stabilised the mean ratio (η) both for new confirmed cases and deaths despite having the earlier cases of COVID-19 and vast quantities of air traffic incoming from neighbouring countries, China, ROK and, of course, the Prince Diamond Cruise Ship outbreak.^19^ But the mean ratio (η) is now again showing a trend of progressing cases which could develop into an alarming situation for Japan. It shows that countries which display a good health security index despite of all their government and individual measures, still struggle to contain the virus and took long time to get stabilised their mean ratio (η) whereas countries which are low in health security index, like Iran, are at massive risk for their entire population to become infected with COVID-19. The mean ratio (η) of new cases for Iran is still progressing and more alarming, the death rate mean ratio (η) is showing increased intensity. Our model predicts that if the world community does not assist the countries with a low health security index the virus will cause thousands of deaths in the coming 30 days. Specifically Iran has UN sanctions in place creating a further complicated situation. Recently Iran has applied to the International Monetary Fund (IMF) for 5 billion USD to fight the COVID-19 pandemic, even Iran secures the loan, they would be unable to get medical supplies as the US sanctions make the any type bank transaction impossible.^27^ Our model estimation is based upon WHO reports, however, according to the international media, the actual numbers of cases and deaths in Iran is much higher than official reports. Even with this available data, the set mean ratio (η), the maximum bound limit is indicating that the entire Iranian population is at increased risk of developing the infection. This is a very alarming finding as Iran borders with countries which are war-torn and this infection can run rampant through those countries populations especially, neighbouring countries like Iraq, Afghanistan, Yemen and to some extent Pakistan. Pakistan has already reported 887 confirmed cases, 6 deaths and 2,923 suspected cases.^28^

These countries need to enhance their precautionary measures by voluntary plus mandated quarantine, stopping mass gatherings, the closure of educational institutes or places of work where the infection has been identified, and the isolation of households, towns, or even cities. There are report that a 1? rise in temperature led to a decrease of the cumulative number of cases by 0.86.^29^ There is hope that a warm summer in this region might help to contain this virus but no change has been seen thus far. Therefore, individual behaviour will be crucial to control the spread of COVID-19. Personal responsibility, rather than government action may make a difference. Early self-isolation, seeking medical advice remotely unless symptoms are severe, and social distancing are key to overcoming this pandemic. Government actions should focus on providing good diagnostic facilities along with providing remotely accessible health advice, together with specialised treatment for those with severe cases of the disease.

As our mathematical model is based on the latest and large scale real-time data and it should provide an additional advantage over the other modelling studies for COVID-19. Secondly, our model was not based solely on mobility data and a large proportion of that data was from the phase of the pandemic when countries had already taken measures and travel bans were in place. The only limitation of our model is that we did not consider the worldwide temperature differences and seasonality of coronavirus transmission. If 2019-nCoV, similar to influenza, has strong seasonality in its transmission, our epidemic forecast might be biased. Furthermore, we did not account for a vaccine to become publicly available to reduce the impact of the pandemic. But even in the absence of a vaccine, human behaviour and the environment itself can alter the likelihood of spread. Hospitals isolate infected people or they may self-quarantine. A further decrease also often occurs as an outbreak matures as people become immune because of previous exposure, reducing the number of susceptible hosts. It has been recently reported that COVID-19 can stay active in aerosol droplets for 3 hours and viable on solid surfaces for 72 hours so new precautionary measures according to this information can reduce viral spread.^11^ Further action might include a ban on paper currency notes, a suspension of all cardboard box packed delivery of items and discouragement of human-to-human contact. By using this model world health agencies need to prepare their AID program in advanced for health resources limited countries before the pandemic cause huge damages.

## Conclusion

It has been found that countries with limited health resources are significantly at high risk and among high health security index countries. Japan has been found more successful in controlling the trend and maintain the pandemic stabilised condition in its country. Countries need to learn lessons from Japan, South Korea and China to get control over COVID 19 pandemic. Other than government restricting on social gatherings and enforcing travel bans to mitigate its spread, individuals also need to show responsibility to avoid useless movements. While countries need to increase testing of suspected cases at large scale to save people lives. Our this model study can be used as baseline to estimate the outbreak size and how much vaccine loads will be required to immunised all infected patients. This model can best provide information to reinforce the forward planning needed to act in the face of this lethal world-wide pandemic.

## Data Availability

Data collected from the daily situation reports of WHO from the 21st of January to the 18th of March 2020, daily reports from the Ministry of Health, Labour and Welfare of Japan, National Health Commission of the People of China and the Korean Centre for Disease Control.

https://www.who.int/emergencies/diseases/novel-coronavirus-2019/situation-reports/

https://www.mhlw.go.jp/index.html

http://www.nhc.gov.cn/xcs/yqfkdt/gzbd_index.shtml

http://www.cdc.go.kr/index.es?sid=a2

## Conflict of Interests

Authors have no conflict of interest

## Funding Source

No special funding for this study.

## Authors Contributions

Muhammad Qasim developed the Idea, collected the data, analysed the data and wrote the manuscript. Waqas Ahmad collected the data, mathematically model the data and developed figures. Minami Yoshida collected data. Maree Gould and Muhammad Yasir reviewed the data and manuscript

## Appendix

1. Data sheet

## Notes

### Competing Interest Statement

The authors have declared no competing interest.

